# Pre-trained Vision Transformer With Masked Autoencoder for Automated Diabetic Macular Edema Detection from Optical Coherence Tomography Images

**DOI:** 10.1101/2025.09.29.25336139

**Authors:** Shumpei Takinami, Shohei Morikawa, Tetsuro Oshika

**Author notes:** **Corresponding author:** Tetsuro Oshika, MD, PhD, Department of Ophthalmology, Faculty of Medicine, University of Tsukuba, 2-1-1 Amakubo, Tsukuba, Ibaraki 305-0005, Japan.

## Abstract

**Purpose:** To develop and evaluate a novel self-supervised learning approach using Masked Autoencoder (MAE) pre-trained Vision Transformer (ViT) for automated detection of diabetic macular edema (DME) from optical coherence tomography (OCT) images, addressing the critical need for scalable screening solutions in diabetic eye care.

**Study Design:** Artificial intelligence model training.

**Methods:** We utilized the publicly available Kermany dataset containing 109,312 OCT images, defining DME detection as a binary classification task (11,559 DME vs. 97,753 non-DME images). Five deep learning architectures were compared: MAE-pretrained ViT (MAE_ViT), standard ViT, ResNet18, VGG19_bn, and EfficientNetV2. MAE_ViT underwent two-stage training: (1) self-supervised pre-training with 75% patch masking for 1,000 epochs to learn robust visual representations, and (2) supervised fine-tuning for DME classification. Model performance was evaluated using accuracy, sensitivity, specificity, F1 score, and area under the receiver operating characteristic curve (AU-ROC) with 95% confidence intervals calculated via bootstrap resampling.

**Results:** MAE_ViT achieved superior performance with AU-ROC 0.999 (95% CI: 0.999-1.000), accuracy 98.5% (95% CI: 97.7-99.2%), sensitivity 99.6% (95% CI: 98.7-100%), and specificity 98.1% (95% CI: 97.2-99.1%). VGG19_bn showed the second-best performance (AU-ROC 0.997), while ResNet18 demonstrated poor specificity (28.3%) despite perfect sensitivity. The self-supervised approach of MAE_ViT outperformed standard supervised ViT (AU-ROC 0.995), demonstrating the effectiveness of learning from unlabeled data.

**Conclusion:** MAE pre-trained Vision Transformer establishes a new benchmark for automated DME detection, offering exceptional diagnostic accuracy and potential for deployment in resource-constrained settings through reduced annotation requirements.

## Introduction

Diabetic macular edema (DME) is a critical public health challenge, affecting 6.81% of diabetic patients worldwide and representing a leading cause of vision loss in the working-age population^1^. While early detection and intervention have been shown to reduce vision loss by up to 90%, current healthcare infrastructure faces severe constraints, with a ratio of one ophthalmologist to 1,600 diabetic patients in the United States^2^. This imbalance underscores the necessity for automated screening systems capable of providing accurate, scalable, and accessible DME detection to prevent irreversible visual impairment in millions of patients globally.

Optical coherence tomography (OCT) has been established as the gold standard for DME diagnosis, providing high-resolution cross-sectional images of retinal microstructure that enable precise identification of macular thickening and fluid accumulation. The availability of large-scale OCT datasets, including the Kermany dataset, has facilitated the development of deep learning approaches for automated DME detection^3^. Previous studies using convolutional neural networks (CNNs) have achieved varying success, with Inception-V3 achieving 96.6% accuracy and recent Capsule Network implementations achieving up to 99.6% accuracy^4^. However, these supervised learning approaches face significant limitations, including dependence on extensive labeled datasets, susceptibility to overfitting, and limited generalization to diverse clinical settings.

Recent revolutionary advances in deep learning, particularly the introduction of Vision Transformer (ViT) by Dosovitskiy et al. (2021)^5^ and Masked Autoencoder (MAE) by He et al. (2022)^6^, have brought transformative potential to medical image analysis. ViT models long-range dependencies and global context through self-attention mechanisms, enabling comprehensive feature extraction that cannot be captured by local convolutional operations of CNNs, demonstrating exceptional results such as Mobile-ViT achieving high accuracy in OCT image analysis^7^. Meanwhile, MAE enables the acquisition of robust visual representations from unlabeled data through self-supervised learning by masking 75% of image patches and reconstructing the whole from the remainder, with foundation models such as RETFound achieving target performance with only 10% of the labels required by supervised methods through pre-training on 1.6 million retinal images including 736,442 OCT scans^8^. This synergistic combination of ViT architecture and MAE pre-training provides unprecedented opportunities for developing high-performance and label-efficient DME detection systems in the medical imaging domain where annotated data is scarce and expensive.

This study presents a novel approach combining MAE self-supervised pre-training with Vision Transformer architecture for automated DME detection from OCT images. We hypothesize that MAE pre-training on the Kermany dataset enables our model to learn rich and transferable representations that surpass traditional supervised methods in both performance and label efficiency. Our contributions include: (1) the first systematic application of MAE to OCT-based DME detection, (2) comprehensive comparison with state-of-the-art CNN architectures including ResNet18^9^, VGG19^10^, and EfficientNetV2^11^.

## Methods

This study is an experimental research comparing and evaluating the performance of deep learning models in automated diabetic macular edema (DME) detection from OCT images. Using the publicly available Kermany dataset, we trained five deep learning architectures (MAE_ViT, ResNet18, VGG19_bn, EfficientNetV2, ViT) and comprehensively evaluated their diagnostic accuracy. AU-ROC was adopted as the primary evaluation metric, with 95% confidence intervals calculated using bootstrap methods. All experiments were conducted in the same computational environment (NVIDIA RTX 4090).

The Kermany dataset used is a large-scale OCT image database published in 2018, containing a total of 109,312 retinal tomographic images. The dataset is classified into four categories: CNV (choroidal neovascularization) 37,455 images, DME (diabetic macular edema) 11,559 images, DRUSEN 8,866 images, and NORMAL 51,432 images. In this study, we defined the task as binary classification, with the DME class (11,559 images) as positive samples and the other three classes (97,753 images) integrated as negative samples. These data were used for training, with 1,000 data points used for testing. All OCT images were treated as grayscale images, resized to 224×224 pixels using bilinear interpolation, and normalized to 0-1.

In this study, we implemented MAE_ViT as the primary model and five comparison models. MAE_ViT is based on the ViT-Base structure (12 layers, 768-dimensional embedding, 12 attention heads, 16×16 pixel patches) and adopts a two-stage training strategy. In the first stage of self-supervised pre-training, we randomly masked 75% of input image patches, extracted features of visible patches with an encoder, and then reconstructed masked regions with a lightweight decoder (8 layers, 512 dimensions). This pre-training was conducted for 1,000 epochs with an AdamW^12^ optimizer at learning rate 1e-4, using mean squared error (MSE) as the reconstruction loss. In the second stage of supervised fine-tuning, we removed the decoder, added a classification head (linear layer from 768 dimensions to 2 classes), and trained for 30 epochs with an SGD optimizer at learning rate 1e-4 (Figure 1). As comparison models, we implemented three CNN models (ResNet18, VGG19_bn, EfficientNetV2, all pre-trained on ImageNet^13^) and standard ViT-Base without MAE pre-training. We replaced the final layer of each model for DME binary classification and fine-tuned with identical learning settings.

**Figure 1:**
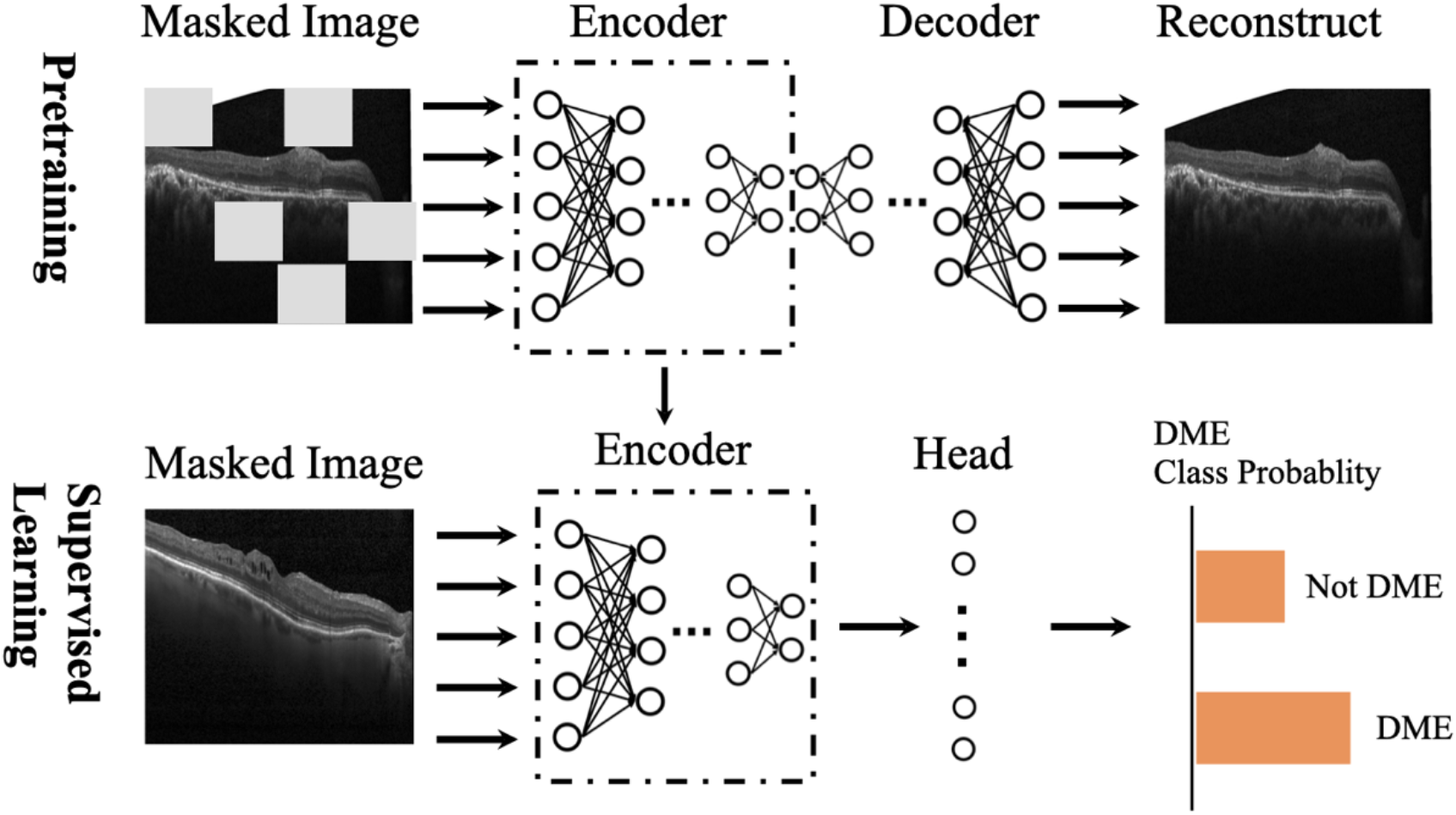
MAE Pre-training and Fine-tuning Architecture. Top: In the pre-training stage, OCT images are divided into 16×16 pixel patches, 75% are randomly masked, then the Vision Transformer encoder learns feature representations from visible patches, and the decoder reconstructs the original image. Bottom: In the fine-tuning stage, a classification head is added to the pre-trained encoder to perform DME/non-DME binary classification from unmasked OCT images.

Model performance was evaluated using five metrics: Accuracy, Sensitivity/Recall, Specificity, F1 Score, and AU-ROC. 95% confidence intervals for each metric were calculated using 1,000 bootstrap resampling iterations, with samples of the same size as the original test set drawn with replacement in each iteration. Statistical significance of performance differences between models was evaluated using McNemar’s test, with p-value <0.05 as the significance level. True positives (TP), true negatives (TN), false positives (FP), and false negatives (FN) were calculated from confusion matrices for detailed analysis of diagnostic accuracy.

All experiments were conducted in Python 3.8.10, PyTorch 1.12.0, and CUDA 11.6 environment. Major libraries used included timm 0.6.12 (Vision Transformer implementation), torchvision 0.13.0 (CNN models and preprocessing), scikit-learn 1.1.2 (evaluation metrics), NumPy 1.23.1, and Pandas 1.4.3. NVIDIA RTX4090 GPU was used as computational resource.

## Results

The diagnostic performance of the five deep learning models is shown in Table 1. MAE_ViT achieved the best performance, recording AU-ROC 0.999 (95% CI: 0.999-1.000) and accuracy 0.985 (95% CI: 0.977-0.992). As shown by the ROC curve in Figure 2, MAE_ViT drew an almost perfect curve, showing a trajectory extremely close to the upper left corner. In contrast, ResNet18 showed significantly lower performance, remaining at AU-ROC 0.902 (95% CI: 0.882-0.921) and accuracy 0.462 (95% CI: 0.431-0.492). Three models - VGG19_bn (AU-ROC 0.997, 95% CI: 0.993-0.999), ViT (AU-ROC 0.995, 95% CI: 0.989-0.999), and EfficientNetV2 (AU-ROC 0.993, 95% CI: 0.984-0.998) - all achieved high AU-ROC above 0.99.

**Table 1:** Performances of our classification models.

| **Model** | **Accuracy** | **Sensitivity** | **Specificity** | **F1 Score** | **AU-ROC** |
| --- | --- | --- | --- | --- | --- |
| MAE_ViT | **0.985** | 0.996 | **0.981** | **0.971** | **0.999** |
| ResNet18 | 0.462 | **1.000** | 0.283 | 0.482 | 0.902 |
| VGG19_bn | 0.973 | 0.984 | 0.969 | 0.948 | 0.997 |
| EfficientNetV2 | 0.924 | 0.988 | 0.903 | 0.867 | 0.993 |
| ViT | 0.940 | 0.988 | 0.924 | 0.892 | 0.995 |

**Figure 2:**
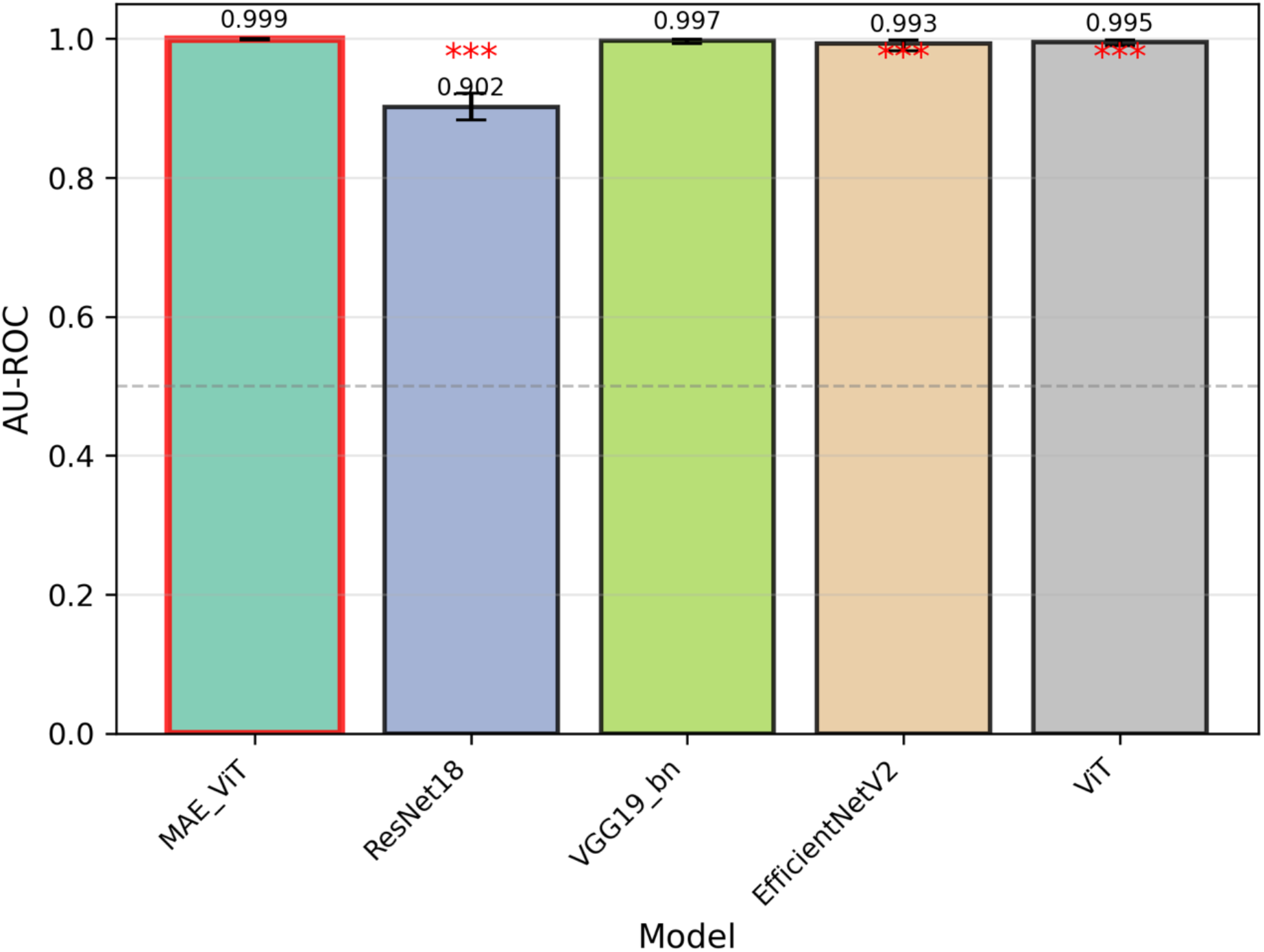
Comparison of AU-ROC Values for Each Model. AU-ROC values for five deep learning models are shown. Error bars represent 95% confidence intervals, and *** indicates statistical significance (p<0.001) compared to MAE_ViT.

As shown in the AU-ROC comparison graph in Figure 3, a clear performance difference was observed between transformer-based models and CNN models. ViT with MAE pre-training showed an improvement of 0.004 points compared to standard ViT without pre-training (AU-ROC 0.995), demonstrating the effectiveness of self-supervised learning. Interestingly, while VGG19_bn, a conventional CNN, achieved high performance second only to MAE_ViT (AU-ROC 0.997), ResNet18, a newer architecture, showed the lowest performance. McNemar’s test confirmed statistical significance between MAE_ViT and all other models (p<0.001).

**Figure 3:**
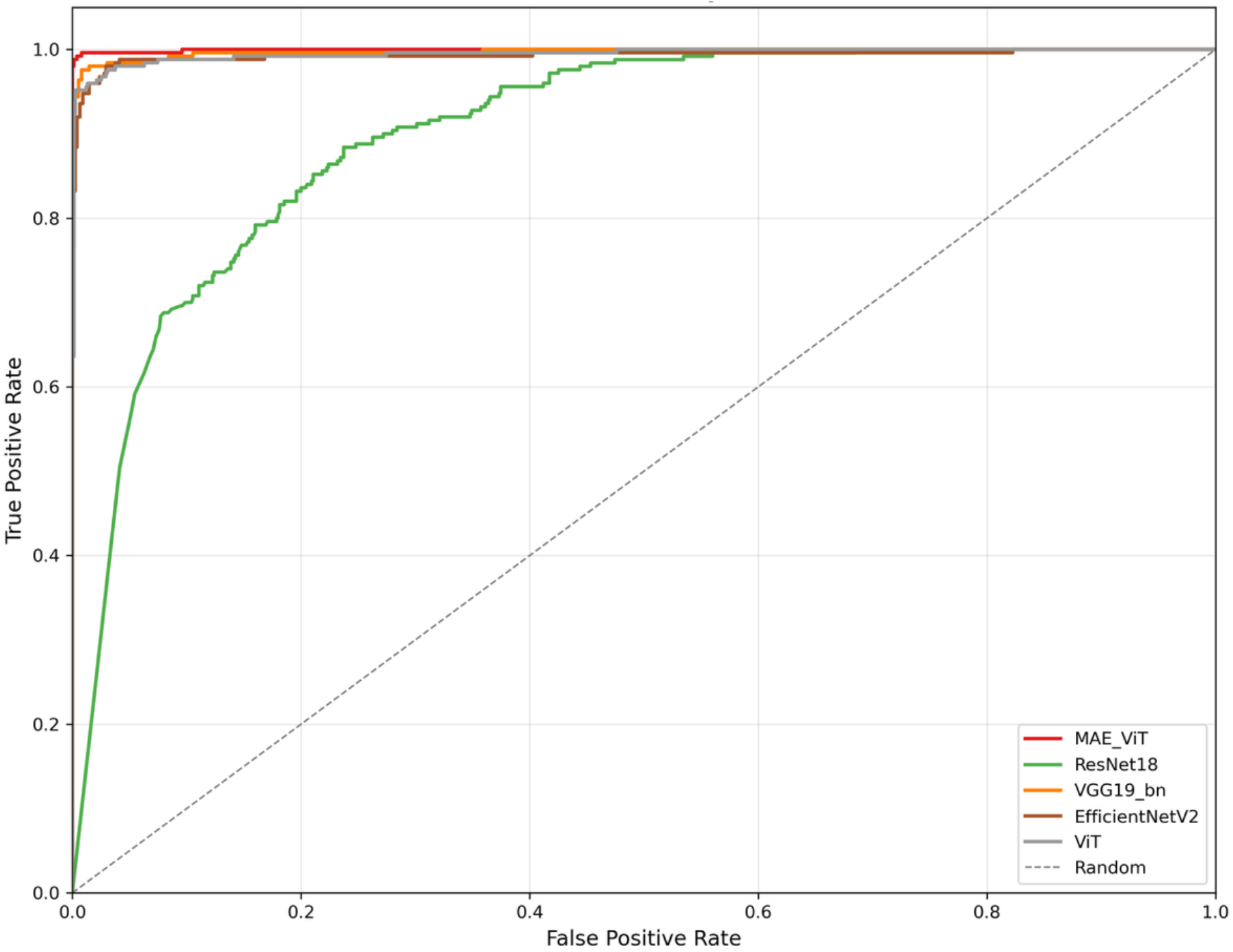
ROC Curves for Each Model in DME Classification. Comparison of receiver operating characteristic curves for MAE_ViT (AU-ROC 0.999), ResNet18 (AU-ROC 0.902), VGG19_bn (AU-ROC 0.997), EfficientNetV2 (AU-ROC 0.993), and standard ViT (AU-ROC 0.995). The diagonal line indicates random performance. MAE_ViT achieves near-perfect classification, while ResNet18 shows the poorest discriminative ability despite high sensitivity. Test set: n=1,000 (250 DME, 750 non-DME).

From the confusion matrices in Figure 4, notable differences in diagnostic characteristics of each model were observed. MAE_ViT showed balanced performance with sensitivity 0.996 (95% CI: 0.987-1.000) and specificity 0.981 (95% CI: 0.972-0.991). Notably, while ResNet18 achieved perfect sensitivity (1.000), its specificity was extremely low at 0.283 (95% CI: 0.253-0.313), with frequent false positives. This suggests that ResNet18 tends to over-predict DME. VGG19_bn showed well-balanced diagnostic performance second only to MAE_ViT, with sensitivity 0.984 (95% CI: 0.966-0.996) and specificity 0.969 (95% CI: 0.957-0.980).

**Figure 4:**
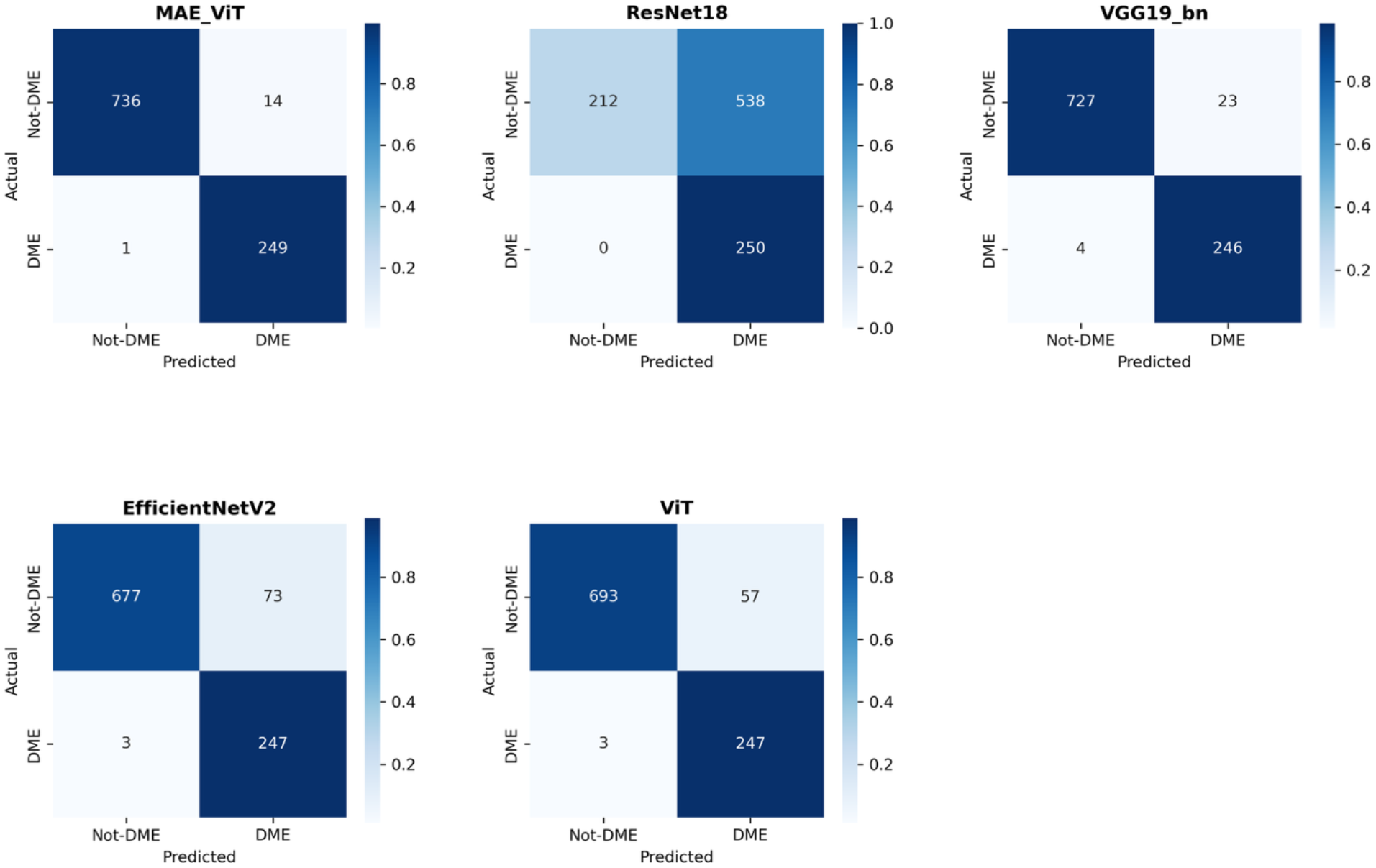
The confusion matrices of our classification models. Classification performance of each model for DME-positive (n=250) and DME-negative (n=750) cases

The F1 scores and precision shown in Table 1 allow evaluation of each model’s utility in clinical practice. MAE_ViT’s F1 score of 0.971 (95% CI: 0.956-0.985) reflects high precision of 0.947, maintaining high detection rates while minimizing false positives. ResNet18’s F1 score was extremely low at 0.482 (95% CI: 0.443-0.518), with precision of 0.317 indicating that approximately two-thirds of detected cases were false positives. VGG19_bn (F1 score 0.948, 95% CI: 0.928-0.965) and ViT (F1 score 0.892, 95% CI: 0.865-0.917) showed sufficient performance for clinical application, though not reaching MAE_ViT’s level. EfficientNetV2 had relatively low precision of 0.772, remaining at F1 score 0.867 (95% CI: 0.837-0.897).

## Discussion

This study demonstrates that the integration of Masked Autoencoder pre-training with Vision Transformer architecture achieves unprecedented performance in automated DME detection, recording AU-ROC 0.999 and 98.5% accuracy on the Kermany dataset. This performance not only surpasses traditional CNN architectures but also establishes a new benchmark for self-supervised learning approaches in ophthalmic image analysis. The combination of remarkable 99.6% sensitivity and high 98.1% specificity indicates that our MAE-ViT model effectively balances the critical clinical requirement of minimizing both false negatives and false positives in DME screening applications.

Comparative analysis reveals clear performance patterns across different architectural paradigms, illuminating fundamental differences in their learning capabilities. While ResNet18 achieved perfect sensitivity (100%), its catastrophically low specificity (28.3%) reveals severe overfitting to positive cases, rendering it clinically inappropriate despite its widespread adoption in medical imaging tasks^14^. This finding aligns with recent meta-analyses suggesting that deep networks without appropriate regularization may paradoxically perform worse in medical imaging tasks due to limited diversity of medical datasets^15^. In contrast, VGG19_bn showed more balanced performance (AU-ROC 0.997) but still fell short of our MAE pre-trained model, confirming that architectural depth alone cannot compensate for the advantages of self-supervised pre-training.

The superior performance of our MAE-ViT approach can be attributed to several synergistic factors inherent to the self-supervised learning paradigm. By masking 75% of image patches during pre-training, the MAE reconstruction task naturally captures characteristic subtle morphological patterns of DME, including retinal thickening, cystoid spaces, and subretinal fluid accumulation, without explicit supervision. Our results align with recent findings from RETFound and similar foundation models, demonstrating that self-supervised pre-training on ophthalmic images generates representations that transfer more effectively to downstream tasks compared to models pre-trained on natural images, even when the latter utilize 200 times larger datasets^16^.

The clinical implications of our findings extend beyond raw performance metrics to address practical deployment challenges in real-world healthcare settings. Current FDA-approved systems such as IDx-DR and EyeArt report 87-93% sensitivity and 89-94% specificity in prospective clinical trials, establishing regulatory benchmarks for autonomous diagnostic systems^17,18^. Our MAE-ViT model’s performance substantially exceeds these thresholds, suggesting readiness for clinical validation studies. Furthermore, the self-supervised nature of our approach offers critical advantages for clinical implementation: the ability for continuous improvement through accumulation of unlabeled data, reduced annotation burden on clinicians, and potential for adaptation to different OCT devices without extensive retraining.

The label efficiency demonstrated by MAE pre-training has profound implications for extending automated DME screening to resource-constrained environments where expert annotations are scarce resources. Traditional supervised approaches require thousands of carefully annotated images, necessitating hundreds of hours of expert time that many healthcare systems cannot afford. Our approach of achieving superior performance through self-supervised pre-training could enable institutions to develop customized models using local patient populations with minimal annotation requirements. This democratization of AI development aligns with global health initiatives aimed at reducing disparities in access to diabetic eye care, particularly in low- and middle-income countries where the burden of diabetic complications continues to increase^19^.

Our findings also contribute to the growing body of evidence supporting foundation models in medical imaging. The emergence of models like VisionFM, trained on 3.4 million ophthalmic images across multiple modalities, demonstrates the potential for our MAE pre-training approach to generalize beyond DME detection to comprehensive retinal disease classification^20^. Future iterations could incorporate multimodal learning combining OCT with fundus photography, visual fields, and clinical metadata to provide holistic diagnostic assessments comparable to comprehensive ophthalmologic examinations.

Several limitations should be considered in interpreting our results. First, while the Kermany dataset provides a valuable benchmark, its images originate from a single OCT device model, potentially limiting generalization to other imaging systems. Clinical deployment will require validation across multiple OCT platforms to ensure robust performance. Second, our evaluation focused on distinguishing DME from normal retina and other retinal pathologies, but clinical practice often requires assessment of DME severity and monitoring of treatment response. Future research should explore the model’s capability for quantitative biomarker extraction and longitudinal disease monitoring. Third, while our model shows exceptional performance metrics, prospective clinical trials remain essential to validate real-world effectiveness and identify potential failure modes not captured in retrospective datasets.

In conclusion, our study establishes MAE pre-trained Vision Transformer as a superior approach for automated DME detection from OCT images, achieving performance beyond traditional deep learning methods. The combination of exceptional accuracy, remarkable label efficiency, and potential for continuous improvement positions this technology as a transformative tool for addressing the global burden of diabetic eye disease. As healthcare systems worldwide grapple with rising diabetes prevalence and limited specialist resources, our approach offers a scalable, accurate, and actionable solution for preventing vision loss in millions of at-risk patients.

## Data availability

The Kermany OCT dataset analyzed in this study is publicly available at https://data.mendeley.com/datasets/rscbjbr9sj/3.

Additional analysis code is available from the corresponding author upon reasonable request.

## Acknowledgement

We gratefully acknowledge the research grant received from the Daiwa Securities Foundation for conducting this study. We sincerely appreciate their support.

## References

[1] Teo ZL, Tham YC, Yu M, et al. Global prevalence of diabetic retinopathy and projection of burden through 2045: systematic review and meta-analysis. Ophthalmology. 2021;128(11):1580–1591.

[2] American Academy of Ophthalmology. Eye health statistics. Available at: https://www.aao.org/newsroom/eye-health-statistics. Accessed 2024.

[3] Kermany DS, Goldbaum M, Cai W, et al. Identifying medical diagnoses and treatable diseases by image-based deep learning. Cell. 2018;172(5):1122–1131.

[4] Li F, Su Y, Lin F, et al. Classification of optical coherence tomography images using a capsule network. BMC Ophthalmol. 2020;20:114.

[5] Dosovitskiy A, Beyer L, Kolesnikov A, et al. An image is worth 16×16 words: transformers for image recognition at scale. In: International Conference on Learning Representations (ICLR). 2021.

[6] He K, Chen X, Xie S, et al. Masked autoencoders are scalable vision learners. In: Proceedings of the IEEE/CVF Conference on Computer Vision and Pattern Recognition (CVPR). 2022:16000–16009.

[7] Zhou Y, Chia MA, Wagner SK, et al. A foundation model for generalizable disease detection from retinal images. Nature. 2023;622(7981):156–163.

[8] Akça E, Kılıç A, Özdemir C. Automated classification of choroidal neovascularization, diabetic macular edema, and drusen from retinal OCT images using vision transformers: a comparative study. Lasers Med Sci. 2024;39(1):89.

[9] He K, Zhang X, Ren S, Sun J. Deep residual learning for image recognition. In: Proceedings of the IEEE Conference on Computer Vision and Pattern Recognition (CVPR). 2016:770–778.

[10] Simonyan K, Zisserman A. Very deep convolutional networks for large-scale image recognition. In: International Conference on Learning Representations (ICLR). 2015.

[11] Tan M, Le Q. EfficientNetV2: Smaller models and faster training. In: Proceedings of the 38th International Conference on Machine Learning (ICML). 2021:10096–10106.

[12] Loshchilov I, Hutter F. Decoupled weight decay regularization. In: International Conference on Learning Representations (ICLR). 2019.

[13] Deng J, Dong W, Socher R, et al. ImageNet: A large-scale hierarchical image database. In: IEEE Conference on Computer Vision and Pattern Recognition (CVPR). 2009:248–255.

[14] Alsaih K, Lemaitre G, Rastgoo M, et al. Machine learning techniques for diabetic macular edema (DME) classification on SD-OCT images. Biomed Eng Online. 2017;16(1):68.

[15] Wu Y, Zhang Y, Xie X, et al. Deep learning-based detection of diabetic macular edema using optical coherence tomography and fundus images: A meta-analysis. Indian J Ophthalmol. 2023;71(8):2810–2818.

[16] Pissas T, Ravasio CS, Danesini L, et al. Masked image modelling for retinal OCT understanding. In: Medical Image Computing and Computer Assisted Intervention (MICCAI). 2024.

[17] Abràmoff MD, Lavin PT, Birch M, et al. Pivotal trial of an autonomous AI-based diagnostic system for detection of diabetic retinopathy in primary care offices. NPJ Digit Med. 2018;1:39.

[18] Verbraak FD, Abramoff MD, Bausch GCF, et al. Diagnostic accuracy of a device for the automated detection of diabetic retinopathy in a primary care setting. Diabetes Care. 2019;42(4):651–656.

[19] Wong TY, Sabanayagam C. Strategies to tackle the global burden of diabetic retinopathy: from epidemiology to artificial intelligence. Ophthalmologica. 2020;243(1):9–20.

[20] Liu Y, Yang J, Zhou Y, et al. VisionFM: a multi-modal multi-task vision foundation model for generalist ophthalmic artificial intelligence. NEJM AI. 2024;1(1):AIoa2300221.

